# Types of Traumatic Experiences in Drug Overdose-Related Deaths: An Exploratory Latent Class Analysis

**DOI:** 10.1101/2023.04.30.23289256

**Authors:** Judy H. Hong, Constanza de Dios, Jessica C. Badawi, Sarah S. Tonkin, Joy M. Schmitz, Consuelo Walss-Bass, Thomas D. Meyer

## Abstract

**Aim:** Drug overdose related-deaths in the US are increasing, with over 100,000 deaths occurring in 2020, an increase of 30% from the previous year and the highest number recorded in a single year. It is widely known that experiences of trauma and substance use very often co-occur, but little is known about the role of trauma in the context of drug overdose-related deaths. Latent class analysis (LCA) was used to classify drug overdose-related deaths based on type of traumatic experiences and individual, social, and substance use characteristics.

**Methods:** Psychological autopsy data were obtained from the University of Texas Health Science Center at Houston (UTHealth) Brain Collection. A total of 31 drug overdose-related deaths collected from January 2016 through March 2022 were included in this study. LCA was used to identify latent factors via experience of four trauma categories (illness/accidents, sexual/interpersonal violence, death/trauma to another, other situations where life was in danger). Generalized linear modeling (GLM) was used to explore differences on demographic, social, substance use, and psychiatric variables between the latent classes in separate models.

**Results:** LCA identified 2 classes: C1 (*n*=12; 39%) was characterized by higher incidence of overall trauma exposure as well as variation in trauma type; C2 (*n*=19; 61%) had lower levels of overall trauma exposure with sexual/interpersonal violence as the most frequent. GLMs indicated that C1 membership was associated with higher incidence of polysubstance use, being married, and having suicidal ideation compared to C2 membership (*p*s<0.05).

**Conclusion:** Among individuals who died by drug overdose, the exploratory LCA identified two distinct subgroups that differed in type of trauma experienced and substance use pattern, the first group having more “typical” characteristics of drug overdoses cases, the other group less typical. This suggests that those at risk of drug overdose may not always exhibit high-risk characteristics.

## Rising Rates of Overdose Deaths in the US

The United States has seen an exponential increase in drug overdose mortality rates in the past 40 years, with over 170 drug overdose deaths daily (Jalal et al., 2018; Seth et al., 2018). During a 12-month period spanning 2020-2021 alone, there were over 100,000 drug-related deaths in the US, an increase of nearly 30% from the previous year and the highest number recorded in a single year (CDC, 2021). Previous attempts to predict drug overdose deaths largely focused on drug use trends, with the rate of deaths due to specific drug types changing over time (e.g., cocaine 2000-2006, prescription opioids 2007-2013). Currently, overdose deaths are being attributed to new drugs, such as fentanyl, and multi- or polysubstance use (Jalal et al., 2018; Jones et al., 2012; Laing et al., 2021), suggesting that efforts to predict and reduce drug overdose deaths should focus broadly on substance use risk factors rather than evolving trends of specific drug use (Jalal et al., 2018; Tori et al., 2020).

## Trauma, Substance Use, and Overdose Deaths

A well-known risk factor for substance use is trauma (Langeland et al., 2002; López-Castro et al., 2015; Schäfer et al., 2014; Schäfer et al., 2009). Those who experience trauma may develop post-traumatic stress disorder (PTSD) or post-traumatic symptoms and turn to drugs and/or alcohol to cope with intrusive thoughts or alterations in arousal; the relationship can also be inversed with substance use preceding traumatic events (Roberts, 2015). Indeed, 20-50% of individuals who seek substance use treatment meet criteria for lifetime diagnosis of PTSD (Van Dam et al., 2010).

Given the association between trauma and substance use, understanding the relationship between specific trauma profiles and substance use patterns may help identify individuals at increased risk for co-occurring trauma and polysubstance use, and consequently, overdose death (Walsh et al., 2018). This is especially relevant given that individuals with trauma history are likely to report multiple traumas of varying types (Hodges et al., 2013; Briere et al., 2008; Kira et al., 2008). Notably, the effect of interpersonal traumas appears to be the most impactful across trauma types (e.g., Kessler et al., 2017; Geoffrion et al., 2022; Chana et al., 2021; Hedtke et al., 2008) resulting in more severe PTSD symptoms, more negative alterations in cognition and mood as well as avoidance (Guina et al., 2018). Furthermore, the intentionality of the trauma is relevant, such that those who experience traumatic events perceived to be caused by deliberate infliction of harm (e.g., assault, war) are more likely to develop PTSD, with symptoms less likely to remit (Santiago et al., 2013). Moreover, type and accumulation of trauma is associated with psychological symptom severity, which has also been connected to substance use. For example, in a female college sample, Krupnick and colleagues (2004) found that students who experienced multiple incidents of sexual/physical abuse had the highest rate of lifetime alcohol and/or drug dependence compared to students who reported other trauma types or singular traumas.

The association between substance use and trauma suggests that individuals with trauma histories may be particularly vulnerable to drug overdose death. Some studies suggest that experiencing a traumatic event induces stress through a regulatory imbalance in the brain that may lead to impulsivity and increased likelihood of engaging in high levels of substance use and misuse (Berk-Clark & Wolf, 2017). This co-occurrence of substance use and impulsivity in traumatized individuals may increase the risk of drug overdose deaths.

## Other Mental Health, Psychosocial, and Sociodemographic Factors

Beyond polysubstance use and trauma, other risk factors have been associated with overdose death, such as other mental health conditions, psychosocial correlates, and sociodemographic status. This has been found for unintentional drug overdose deaths (Martins et al., 2015), intentional drug overdose deaths (suicides; Bohnert & Ilgen, 2019), and across various substances, both prescription (Brady et al., 2017) and illicit (Yarborough et al, 2016). Although some risk factors for overdose deaths are more established, others such as trauma type require more investigation. In this exploratory study, we aim to classify drug overdose-related deaths based on types of traumatic experiences and individual, social, and substance use characteristics.

## Method

### Study Setting and Procedures

The present study utilized data collected as part of the UTHealth Psychological Autopsy Interview Schedule (UTH-PAIS) validation study (Meyer et al., 2022). Following institutional review board approval and consent for donation by the next-of-kin, postmortem brains were obtained from January 2016 through March 2022 by The University of Texas Health Science Center at Houston (UTHealth) Brain Collection in collaboration with the Harris County Institute of Forensic Science. For each deceased individual, we obtained demographic information as well as autopsy and toxicology reports. A trained research assistant administered the UTH-PAIS by telephone with the decedent’s next-of-kin at least 6 weeks after the decedent’s tissue donation. The UTH-PAIS collected sociodemographic information, presence of psychiatric clinical phenotypes (e.g., depression, substance use disorder), history of alcohol and drug use, and family history of mental health problems. Interrater reliability ranged from good to excellent; details are described elsewhere (Meyer et al., 2022). A consensus of the decedent’s diagnoses was determined by three trained clinicians (i.e., a psychologist and two psychiatrists), including a review of all available information, including the psychological autopsy, medical records, and medical examiner’s autopsy report.

To be included in the current study, autopsy and toxicology reports had to indicate that the cause of death was due to a drug toxicity, and the UTH-PAIS had to indicate the presence of at least one trauma event. Of 146 screened for eligibility, a total of 31 met criteria to be included in the final sample. See Table 1 for subject (i.e., decedent) demographic information.

**Table 1.**
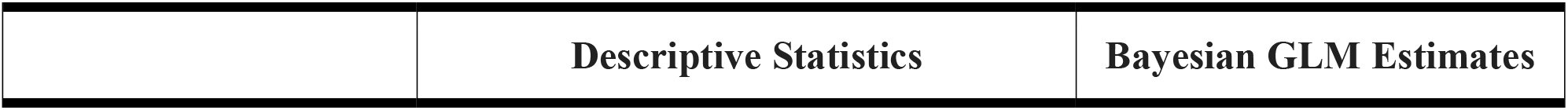
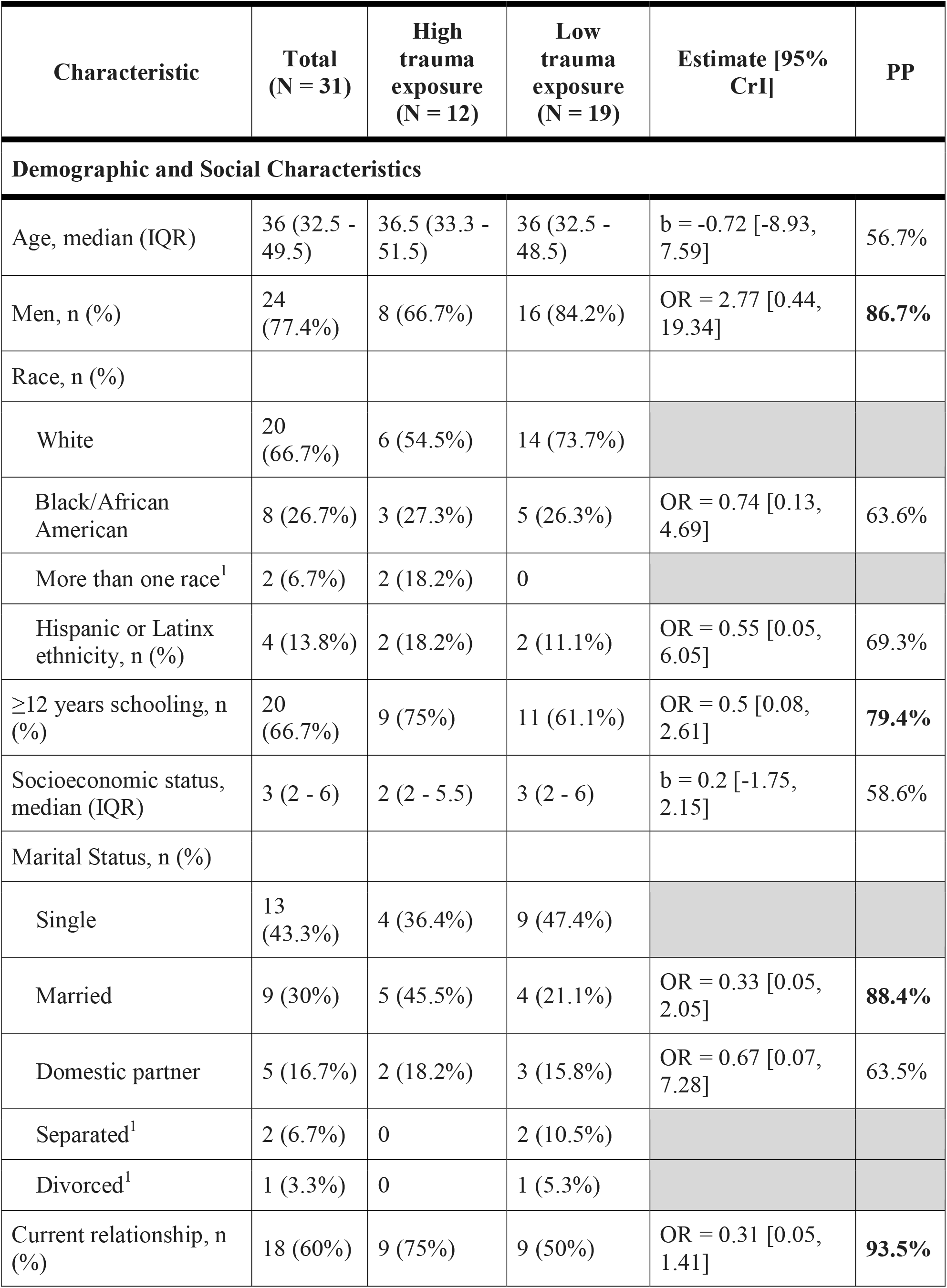
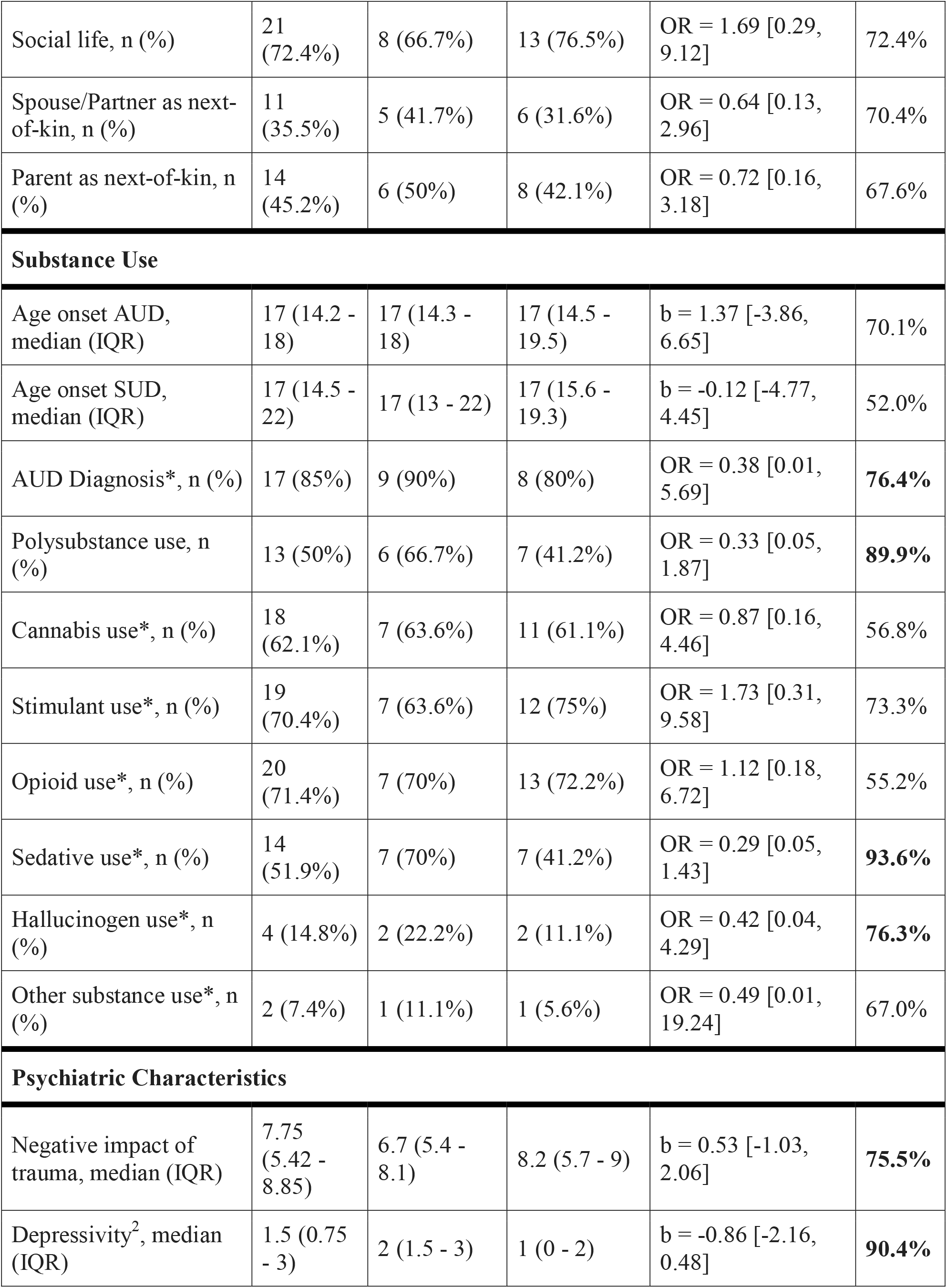
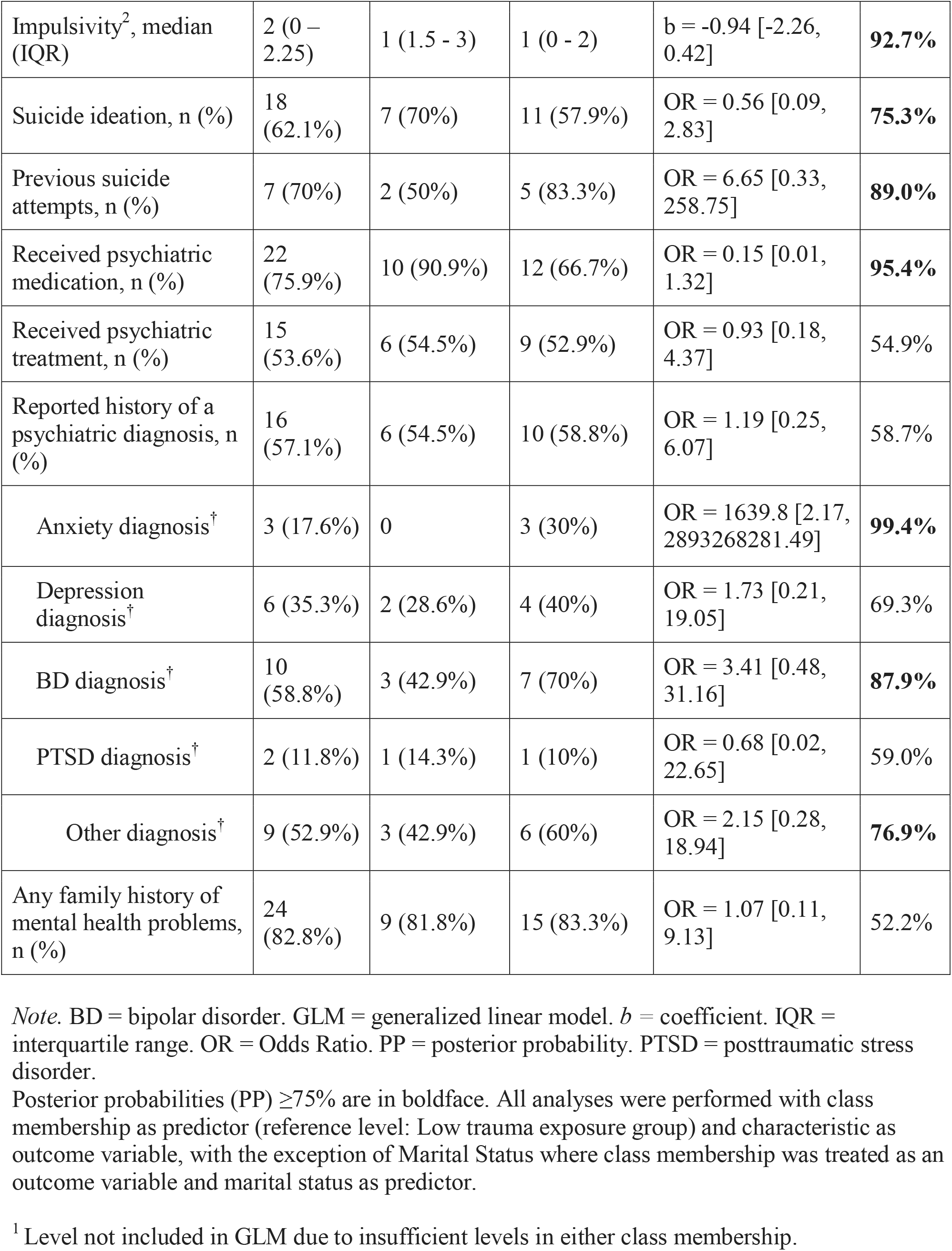
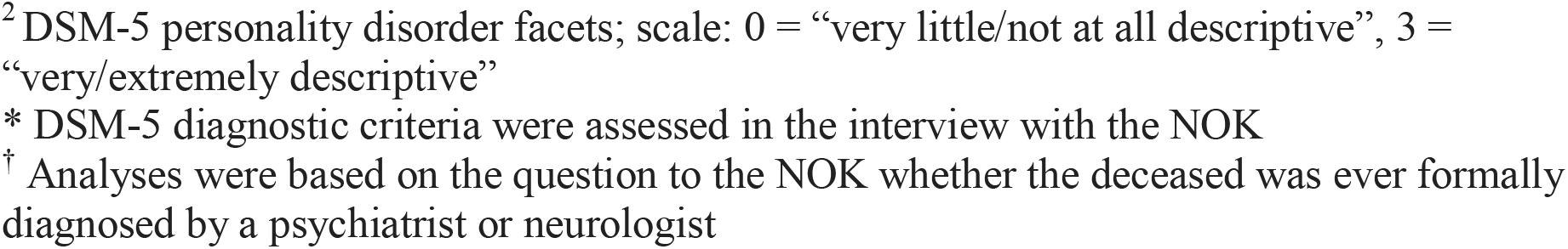
Demographic, Social, Substance Use, and Psychiatric Characteristics by Class Membership

### Measures/Variables of Interest

#### Trauma Categories

The set of nine trauma items were grouped into broader categories based on the Life Events Checklist for DSM-5 (LEC-5, Weathers et al., 2013) as follows: Category 1) Illness and accidents (consisting of Illness and Accident); Category 2) Sexual or interpersonal violence (Sexually abused, Physical force/weapon use, Slapped/beaten/attacked, and Threatened with weapon); Category 3) Death or trauma to another person (Present when another was killed/injured/assaulted and Family/partner/close friend died of accident/homicide/suicide); and Category 4) Any other situation where the decedent’s life was in danger (e.g., military combat, war zone).

Each category was dichotomous, such that an individual was coded as “yes” for a category if they experienced at least one of its constituent items, e.g., an individual who experienced items 2 (accident) and 3 (physical force/weapon use) would be coded as “yes” for Categories 1 and 2. Trauma categories were not mutually exclusive.

Negative impact of trauma was assessed by asking the next-of-kin (NOK), “On a scale from 1 (‘not at all’) to 10 (‘very much’), how much did this negatively affect [decedent’s] well-being?”. This was asked for each of the nine trauma items if they were endorsed, as experienced by the decedent. For each trauma category calculated, if a category comprised more than one trauma item and a decedent was reported to have experienced more than one item in a given category, their trauma impact score was averaged across items, producing one average trauma impact score per category for each decedent. This was done so that if a person experienced traumas across different categories, they could all be included.

#### Demographic and Social Variables

The UTH-PAIS assessed age, gender, race, ethnicity, education, socioeconomic status, and marital status of decedents, including questions about social support networks and whether decedent was in a current relationship. Relationship of NOK to the decedent was characterized as “partner” if NOK was identified as wife, husband, spouse, or partner. Likewise, “parent” was characterized by both parents, mother, or father.

#### Substance Use Variables

Alcohol and substance use problems were assessed by asking NOK “Did you ever have the impression that [he/she] has had a substance use problem” with follow-up questions based on DSM-5 criteria for an alcohol use disorder (AUD) or other substance (cannabis, hallucinogens, opioids, sedatives, stimulants, inhalants, PCP) or polysubstance (use of more than three different substance classes) disorder. Age-of-onset of problematic alcohol use and substance use problems were also assessed.

#### Psychiatric Variables

For the present analyses psychiatric variables assessed were dichotomously reported as yes/no to the question “Did he/she ever see and get a diagnosis from a neurologist or psychiatrist?” These included anxiety, depression, bipolar disorder, PTSD, psychosis, OCD, eating disorder, or “other diagnosis” that could be specified.

Using the DSM-5 personality disorder facets, we also assessed ‘depressivity’ and ‘impulsivity’ via Likert scale asking NOK how well the characteristic described the decedent. For depressivity, the UTH-PAIS assessed the personality disorder traits as outlined in DSM-5 (“Frequent feelings of being sad, down, and/or hopeless; difficulty recovering from such moods; pessimism about the future; frequent feelings of shame; thoughts of suicide and suicidal behavior”, p. 779) and impulsivity (“Acting on the spur of the moment without a plan or consideration of outcomes; difficulty establishing and following plans”) using ratings from 0 (“Very little or not at all descriptive”) to 3 (“Very or Extremely Descriptive”). Suicide ideation (“Has he/she/name ever thought about suicide or even tried to commit suicide?”) and previous suicide attempts (“Had he/she made a suicide attempt”) were adapted from the Columbia-Suicide Severity Rating Scale (Posner et al., 2011). History of psychiatric medication (“Have they ever been prescribed or used any medication to deal with stress, emotional or mental health problems”), treatment other than medication (“Any other treatment like ECT, light therapy, or psychological treatments?”), and decedents’ family history of mental health problems (“Is there any family history of mental health problems including suicidality and alcohol or drug abuse?”) were also assessed.

### Data Analytic Strategy

#### Latent Class Analysis

Latent profiles of trauma experience were identified using latent class analysis (LCA). The four trauma categories were fed as observed variables. We first estimated a model of class size *k* = 1, then added classes until we reached *k* = 4. Each class size k was tested in 10 repetitions so that each repetition started with different random values of class-conditional probabilities. The algorithm returns the repetition with the greatest log-likelihood. This was repeated for each test *k*.

The best model with *k* number of classes was determined based on model fit via Bayesian Information Criterion (BIC), with lower BIC denoting better fit (Nylund, Asparouhov, & Muthén, 2007). Model quality was assessed using entropy, (a measure akin to R-squared) with a value > 0.8 and closest to 1 indicating the most accurate model (Celeux & Soromenho, 1996). As all N = 32 individuals had at least one trauma, no missing data for the input variables was observed, hence all trauma categories and all observations were included in the LCA.

After identifying the best class model, each subject was assigned to the predicted class based on their posterior class membership probabilities. We sought to group subjects based solely on their patterns across the four broad trauma categories, hence no covariates were included in the LCA.

#### Generalized Linear Modeling

Generalized linear modeling (GLM) was used to explore differences on demographic, social, substance use, and psychiatric variables between the classes identified by the LCA. Binary variables were modeled via the binomial distribution, akin to logistic regression.

Parameter estimates for the predictor effects on the outcome in logistic regression were exponentiated to provide odds ratios (OR). Continuous variables were modeled via a student distribution. Each variable of interest was examined in its own separate model. Missing data or “unknown” responses for a variable were excluded only when modeling a particular variable. (See Table S2 for number of missing observations per variable.)

Bayesian inference was used in parameter estimates, as it provides the ability to evaluate the alternative hypothesis for each model (i.e., that differences between the classes exist). The median and 95% credible intervals (CrI) of the posterior distribution were used to provide a point estimate and corresponding range of uncertainty for the magnitude of the predictor effect.

Convergence diagnostics (i.e., Rhat) and effective sample size were examined to ensure satisfaction of Bayesian modeling assumptions. Graphical plots of posterior predictive distributions were visually inspected to verify that the observed distribution of each outcome fell within the range of distributions produced by 1000 replications drawn from the posterior predictive distributions of the outcome. All of the statistical modeling satisfied each of these assumptions.

Probabilities that the parameter estimates exist were quantified using posterior probability (PP), or the extent to which the density of the posterior distribution was less than or greater than zero. Strength of evidence for the alternative hypothesis in each model was based on heuristics for varying PP thresholds described in the literature (Jeffreys, 1961; Lee & Wagenmakers, 2013): (1) no evidence (PP = 50%); (2) anecdotal evidence (51% ≤ PP ≤ 74%); (3) moderate evidence (75% ≤ PP ≤ 90%); (4) strong evidence (91% ≤ PP ≤ 96%); (5) very strong evidence (97% ≤ PP ≤ 99%); and (6) extreme evidence (PP > 99%). The PP ≥ 75% was chosen as a minimum level of evidence in favor of considering effects potentially worthy of future investigation.

All analyses were conducted on the R statistical computing environment (R Core Team, 2022). LCA was performed using the poLCA package (Linzer & Lewis, 2011), and Bayesian GLM via the brms package (Bürkner, 2017).

## Results

### Latent Profiles of Trauma Experience

The best model suggested by BIC was class size *k* = 2, showing adequate entropy, so we selected the 2-class model (Table S1 shows LCA fit statistics for different class models until size 4 classes).

Figure 1 shows conditional probabilities of the profiles of the two latent classes. Class 1 generally had higher levels of trauma exposure as indicated by reporting at least one of the four trauma categories, with illness/accidents (Cat 1), sexual/interpersonal violence (Cat 2) and experiencing death/trauma to another person (Cat 3) experienced by all (100%) class members, thus Class 1 was labeled the “high trauma exposure” class. Fewer members of Class 2 experienced the trauma categories, and none experienced other situations where the decedent’s life was in danger (Cat 4), thus this group was labeled the “low trauma exposure” class.

**Figure 1.**
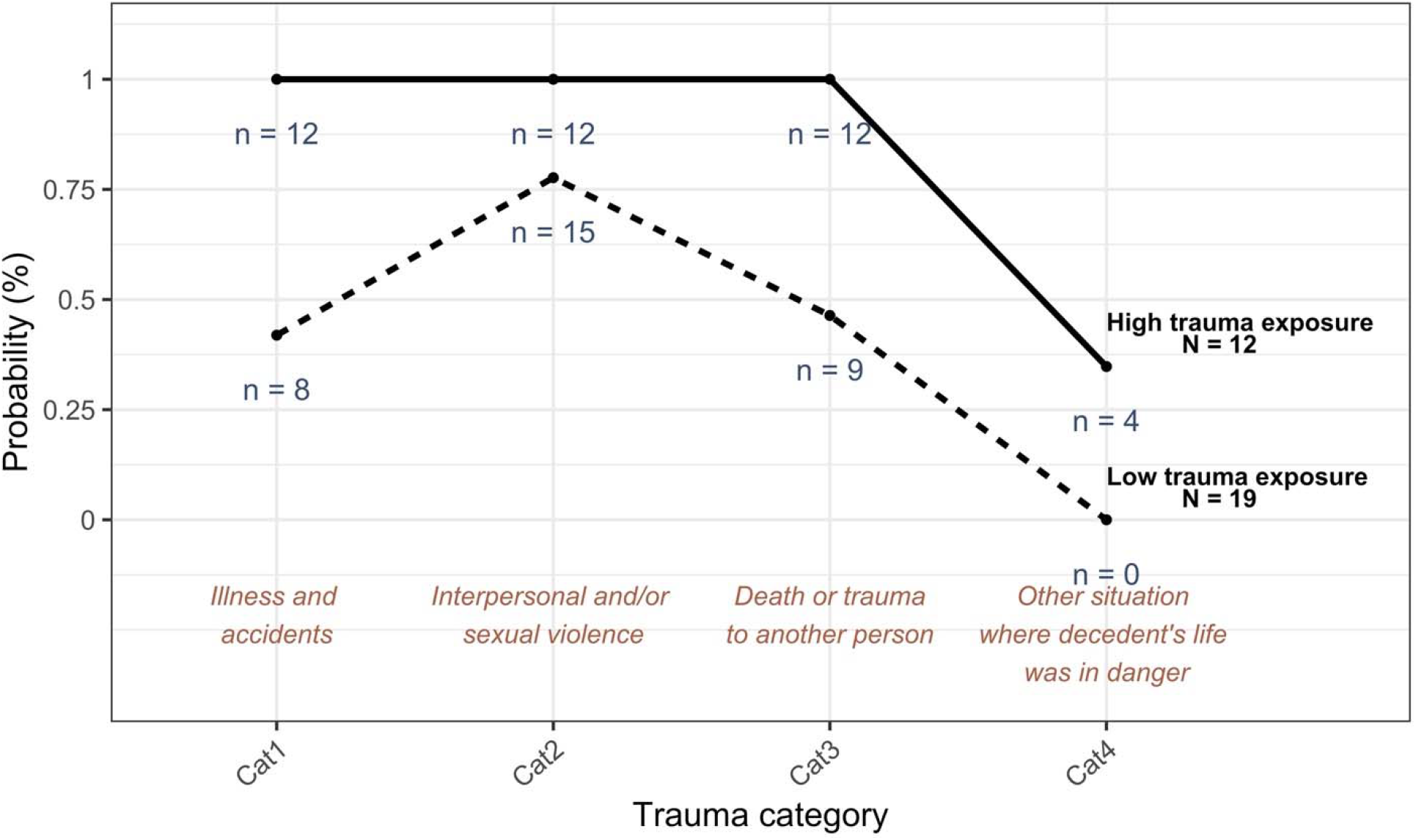
Count and Probability of Trauma Categories in each Latent Class Note: Categories are not mutually exclusive.

### Outcomes by Class Membership

Descriptive characteristics of overall decedent sample (N = 31) is presented in Table 1. Notably, this sample experienced more traumas related to interpersonal and/or sexual violence than any other trauma type: 100% in the high trauma exposure class and 77.7% in the low trauma exposure class.

Comparisons of the two classes via GLM on the demographic, social, substance use, and psychiatric characteristics are also displayed in Table 1. All models except marital status used class membership as predictor (2 levels; high trauma exposure as reference level) and characteristic as outcome. In the case of marital status, a categorical unordered variable, the equation structure was flipped so that marital status was used as predictor (3 levels; “Single” as reference level), and class membership was used as an outcome, producing a logistic regression giving odds ratios of a marital status being in the low trauma exposure group.

### Demographics

The sample had a median age of 36 (interquartile range = 32.5 - 49.5; range: 19-62). There was no evidence for differences in age, race, ethnicity, or socioeconomic status (PP < 75%). There was moderate to strong evidence, however, for differences in gender distribution (low trauma exposure associated with higher incidence of men), education (low trauma exposure associated with lower incidence of completing high school or equivalent), and relationship status (low trauma exposure associated with lower odds being in current relationship). There was also an association with marital status, such that being married was associated with lower odds of being in the low trauma exposure group.

### Substance Use

Regarding substance use, no differences were observed with respect to age of onset for substance or alcohol use, cannabis use, stimulant use, opioid use, or other substance use. Low trauma exposure was associated with lower odds of AUD diagnosis. Low trauma exposure was also associated with lower odds of polysubstance use, as well as use of sedatives and hallucinogens, compared to the high trauma exposure group. In summary, those with high trauma exposure were more likely to show problematic drinking and use of multiple substances.

### Psychiatric Variables

The low trauma exposure group was associated with higher trauma impact scores (measuring negative impact of trauma) compared to the high trauma exposure group (*b* = 0.53). With respect to other psychiatric variables, the low trauma exposure group was associated with lower impulsivity and depressivity compared to the high trauma exposure group. Low trauma exposure was also associated with lower odds of suicide ideation, but higher odds of previous suicide attempts than the high trauma exposure group. The low trauma exposure group was less likely to have received psychiatric medication, but no difference in odds of having received other psychiatric treatment. Low trauma exposure was associated with diagnoses of anxiety, bipolar disorder, or “other” mental disorder diagnosis. No differences were observed in rates of reported history of major depression diagnoses, PTSD diagnoses, or general family history of mental health problems.

## Discussion

In this exploratory study, we used latent class analysis to classify drug overdose-related deaths based on types of traumatic experiences and individual, social, and substance use characteristics. Our overall sample reflected characteristics of drug overdose deaths in other studies: mostly men (Hedegaard et al., 2022) in the 35-44 age range (Hedegaard et al., 2022), who completed high school (Kedia, 2020), with trauma histories (Clark et al., 2022), poor mental health (Kedia, 2020), problems with substance use (Kariisa et al., 2022), past suicidality (Bohnert et al., 2010), that died of unintentional drug overdoses (Hedegaard et al., 2022). Of particular importance, this sample experienced interpersonal trauma at higher rates than other types of trauma. As reflected in the trauma literature, compared to non-interpersonal traumas, interpersonal traumas are more highly associated with PTSD symptoms and predict coping through use of substances (Ullman, 2013).

Previous studies highlight the “typical” drug overdose decedent profile as polysubstance users who experience a large number and varied types of trauma, consistent with the Class 1 “high trauma exposure” group here (Peppin et al., 2020; CDC, 2022; Gilbert et al., 2022). For this group, NOK were more likely to report that the deceased thought about suicide, but were less likely to have made a previous attempt. The personality facets *impulsivity* and *depressivity* were associated with Class 1 membership. While it cannot be determined if the traumatic events preceded or followed the use of substances, or if the overdose death in these cases was accidental or intentional, our findings highlight the interactive effects of trauma and substance use in drug overdose risk.

A larger percentage of cases in our sample (61%), however, comprised the Class 2 “low trauma exposure” group with a distinctly different profile. For this group, the most probable trauma category was sexual and interpersonal violence. As where this group had fewer and less varied traumas than Class 1, they were more likely to have higher trauma impact scores, reflecting greater negative consequences on the individual’s well-being. A diagnosis of alcohol use disorder (meeting DSM-5 criteria) and polysubstance use was less likely in this low trauma exposure group.

For those with impactful interpersonal and/or sexual traumas, the explanatory mechanisms from trauma to substance use may be different compared to other traumas. In a study of young adults, betrayal trauma (i.e., interpersonal violence, that occurred in childhood or adulthood, perpetrated by someone the victim relies on for support; Freyd, 1996) experienced prior to age 18 was associated with problematic substance use via post-traumatic stress and two pathways: self-destructiveness and difficulty discerning/heeding risk (Delker & Freyd, 2014).

This was explained through theories of self-blame and self-preservation. Delker & Freyd (2014) postulated that since the victim may depend on the perpetrator for material or emotional support, such as when a child depends on a caregiver, it may benefit the victim to “remain fully or partially unaware of the abuse” for the purposes of self-preservation and survival. With continued unawareness, the victim may begin to have difficulties discerning or heeding risk.

Furthermore, victim self-blame has been well-documented in the trauma literature (e.g., Frazier, 1990; Janoff-Bulman, 1979) – in attempts to gain a sense of control and understanding of the traumatic event, those who have experienced trauma may blame themselves for the occurrence with the belief that if they take different actions in the future, they could prevent the experience from happening again. This self-blame on the part of the victim may lead to a lack of concern for their own well-being, resulting in self-destructive behavior. For our low trauma exposure group, this self-destructiveness and difficulties discerning/heeding risk may have increased chances for drug overdose. It is noteworthy that members in this group were more likely to be single men, consistent with prior studies reporting a relationship between single relationship status (Aram et al., 2020) and male gender (Hedegaard et al., 2022). That this group also had higher odds of having an anxiety or bipolar disorder and previous suicide attempts, underscores the impact of comorbid psychiatric disorders on drug overdose deaths.

### Limitations, Future Directions, and Conclusions

Our study was not without limitations. Due to the focus on drug overdose cases with psychological autopsies collected as part of a brain collection, our sample was small. However, our sample was representative of those receiving medical autopsies (Meyer et al., 2022).

Additionally, there was only inquiry about whether a trauma type occurred, and not whether there were multiple incidents of the same trauma type. We also did not have chronological information in case of multiple substance use, so we could not determine if it preceded or followed a traumatic event. An inherent limitation in psychological autopsy studies is reliance on next-of-kin to assess characteristics of the deceased. However, as a strength, the new UTH-PAIS, used in this study, has demonstrated good to excellent reliability for psychiatric diagnostic and dimensional characteristics by using standardized assessment instruments (Meyer et al., 2022).

This study is the first to classify drug overdose-related deaths based on type of traumatic experiences and individual, social, and substance use characteristics, which could greatly impact clinical interventions for preventing overdose-related deaths. Specifically, clinicians and other providers can use these findings to pay particular attention to those who have experienced impactful interpersonal traumas and may be using substances to cope. Although an individual may not have a long, varied history of traumatic events or polysubstance use, they may still be at high risk for drug overdose.

As this is an exploratory study, we present suggestions for future research questions:

1. Are the mediating pathways from trauma to substance use different for those with higher versus lower trauma exposure? If so, what are the explanatory factors?
2. What is the chronological order of events from trauma to substance use to overdose for those with lower trauma exposure versus higher trauma exposure?
3. How are the substance use patterns different for those with higher versus lower trauma exposure?

Overall, our findings suggest that it may be informative to identify clusters and different trajectories of drug overdose decedents who experienced traumatic events. These trajectories do not always follow a typical path and more discernment of those who appear lower risk may decrease drug overdose rates.

## Data Availability

All data produced in the present study are available upon reasonable request to the authors.

