## Supplemental Tables S1 and Table S2 for "Types of Traumatic Experiences in Drug Overdose-Related Deaths: An Exploratory Latent Class Analysis"

**Table S1**

*Table of Fit Statistics to Different LCA Class Solutions (Supplement)*

| **Model size (number of classes)** | **LL** | **BIC** | **aBIC** | **cAIC** | **Likelihood ratio** | **Entropy** |
| --- | --- | --- | --- | --- | --- | --- |
| 1 | -65.21 | 144.29 | 131.92 | 148.39 | 14.7 | - |
| 2 | -60.50 | 151.90 | 123.86 | 160.90 | 5.01 | 0.61 |
| 3 | -57.99 | 164.06 | 120.45 | 178.06 | 0.000000002 | 0.97 |
| 4 | -57.99 | 181.23 | 122.04 | 200.23 | 0.0000000008 | 0.56 |

*Note:* LL = log-likelihood; BIC = Bayesian information criterion; aBIC = sample-size adjusted BIC; CAIC = consistent Akaike information criterion.

**Table S2**

*Number of Participants excluded per Variable due to Missingness*

| **Characteristic** | **n Excluded** |
| --- | --- |
| Age | 0 |
| Men | 0 |
| Race | 3 |
| Hispanic or Latinx ethnicity | 2 |
| ≥12 years schooling | 1 |
| Socioeconomic status | 1 |
| Marital Status | 4 |
| Current relationship | 1 |
| Social life | 2 |
| Spouse/Partner as next-of-kin | 0 |
| Parent as next-of-kin | 0 |
| Age onset AUD | 17 |
| Age onset SUD | 10 |
| AUD Diagnosis | 11 |
| Polysubstance use | 5 |
| Cannabis use | 2 |
| Stimulant use | 4 |
| Opioid use | 3 |
| Sedative use | 4 |
| Hallucinogen use | 4 |
| Other substance use | 4 |
| Negative impact of trauma | 0 |
| Depressivity | 15 |
| Impulsivity | 15 |
| Suicide ideation | 2 |
| Previous suicide attempts | 21 |
| Received psychiatric medication | 2 |
| Received psychiatric treatment | 3 |
| Psychiatric diagnosis | 3 |
| Any family history of mental health problems | 2 |
